# Four methods for estimating hepatitis C incidence using extant testing data

**DOI:** 10.1101/2025.10.08.25337617

**Authors:** William J. McFarlane, Jennifer A. Flemming, Susan B. Brogly, Yingwei Peng

**Affiliations:** Department of Public Health Sciences, Queen’s University, Kingston, ON, Canada; ICES Queen’s, Kingston, ON, Canada; Department of Medicine, Queen’s University, Kingston, ON, Canada; Department of Surgery, Queen’s University, Kingston, ON, Canada

**Author notes:** Corresponding author: William McFarlane,. Mailing Address: 10 Stuart St., Kingston, ON, Canada, K7L 3N6.

**Keywords:** Hepatitis C, Incidence, Routinely Collected Health Data, Administrative Health Data, Epidemiologic Methods

## Abstract

**Background:** Extant hepatitis C virus (HCV) antibody (Ab) and ribonucleic acid (RNA) test results are widely used to estimate HCV incidence, but the impact of cohort specification and case definition on validity and generalizability is poorly understood.

**Methods:** Using databases linked at ICES, a cohort of 15.8 million Ontarians aged 18–80 between 1999 and 2018 was used to estimate annual HCV incidence using four methods: the population-based method defined incidence as new annual HCV cases divided by population size estimates; the test-negative method defined eligibility at first negative test; the RNA-based method prioritized specificity by requiring RNA+ tests; and the antibody-inclusive method prioritized sensitivity by including all Ab+ tests. Method assumptions, potential misclassification, and sensitivity to analytic choices were assessed.

**Results:** RNA-based estimates were lowest and fluctuated around 30 cases per 100,000 person-years, while population-based and antibody-inclusive estimates were 1.5-fold higher, and test-negative estimates were 7.9-fold higher. Population-based estimates were influenced by changes in the HCV case definition used in Ontario from 1999-2018. The test-negative cohort had a high prevalence of HIV and substance use disorder, limiting generalizability of HCV incidence estimates. RNA-based estimates likely underestimated HCV incidence because 22% of Ab+ tests were unconfirmed by RNA testing, while antibody-inclusive estimates likely overestimated HCV incidence by assuming all unconfirmed Ab+ tests were true cases.

**Conclusion:** These findings illustrate the influence of cohort definition and HCV case definition when estimating HCV incidence using extant testing data, which will support more accurate monitoring of HCV burden and progress toward elimination.

## Introduction

Extant hepatitis C virus antibody (HCV Ab) and HCV ribonucleic acid (RNA) test results conducted in public laboratories are used to estimate HCV incidence in many regions, including Australia, Canada, and the USA.[1–3] This data source is efficient but does not clearly differentiate prevalent and incident HCV infections, so the potential for misclassification of HCV status is high. There are different approaches for defining HCV-susceptibility and incident HCV infection when using extant testing data, and the impact of these methodological decisions on misclassification and generalizability is poorly understood.

The simplest way to use extant testing data to estimate HCV incidence is a ratio of the number of reported HCV infections to the estimated population size. Issues with this method include changes in HCV case definitions over time, and difficulty differentiating incident and prevalent HCV infections.[4] Chronic HCV infection can persist for decades without symptoms, making it difficult to determine when infection occurred.[5] Incident and prevalent infections are better differentiated using a cohort that defines eligibility based on first testing HCV negative (HCV-).[6, 7] Generalizability of HCV incidence estimates may be limited when using this approach because HCV screening is risk-based in Ontario; test-negative cohorts therefore tend to favor patients with a high risk of HCV such as individuals infected with human immunodeficiency virus (HIV) or with a history of injection drug use (IDU).[8, 9]

HCV incidence can also be estimated using a sample of the general population, by considering individuals HCV- if they have no history of an HCV positive (HCV+) test result. This design improves generalizability compared to the test-negative method, but is more likely to misclassify prevalent HCV infections as incident. One way to increase the specificity of HCV classification is to only consider positive HCV RNA tests (RNA+) when defining incident HCV infection. The HCV testing cascade begins with an HCV Ab test (measuring lifetime HCV exposure); those testing HCV Ab positive (Ab+) should then receive a confirmatory HCV RNA test (measuring active HCV infection).[8] However, up to 44% of individuals that test HCV Ab+ do not receive a confirmatory HCV RNA test (i.e., an unconfirmed HCV Ab+ result).[10] HCV RNA testing often requires an additional blood draw following an HCV Ab+ test result, and unconfirmed HCV Ab+ tests may occur because patients don’t attend RNA testing appointments or are unavailable for scheduling.[8]

It is possible that many individuals with unconfirmed Ab+ test results were actively infected with HCV at the time they tested Ab+, because 75 to 85% of untreated HCV infections become chronic and RNA testing is a prerequisite for HCV treatment.[5, 8, 11] Limiting the definition of HCV infection to RNA+ test results would therefore prioritizes specificity, but would likely underestimate HCV incidence because some of the unconfirmed Ab+ tests came from individuals who would have tested HCV RNA+ (if tested).[10] Conversely, sensitivity can be maximized by defining incident HCV infection as an RNA+ test or an individual’s first recorded Ab+ test result. This would include all unconfirmed Ab+ tests as cases, likely overestimating HCV incidence because not all of these individuals would have tested HCV RNA+ (if tested).

Extant HCV testing data are an efficient data source for estimating HCV incidence, with data repositories for many regions including the integrated Public Health Information System for Ontario, and the National Notifiable Diseases Surveillance System for the USA.[12, 13] The strengths and limitations of methods available for estimating HCV incidence using extant testing data are poorly understood. Therefore, the objective of this study was to use extant HCV testing data to estimate the annual incidence of HCV from 1999 to 2018 in Ontario using four different methods, and comment on the generalizability and potential biases for each method.

## Data and methods

### Data sources

The study cohort included all universal access Ontario Health Insurance Plan (OHIP) beneficiaries aged 18 – 80 from 1999 to 2018. This follow-up period was chosen based on availability of HCV testing data. The follow-up period was January 1^st^, 1999 to December 31^st^, 2018. Time zero for each individual was defined as the date they became eligible for follow-up if this occurred between 1999 and 2018, or January 1999 if they met the eligibility criteria prior to 1999. The maximum follow-up date was December 31^st^, 2018, an individual’s death date, the date they turned 81, or the date they became ineligible for OHIP.

HCV Ab and HCV RNA laboratory test results from 1999 to 2018 were obtained from Public Health Ontario (PHO). PHO has 11 laboratories across Ontario, which provide services (e.g., HCV testing) to clinicians in primary care, hospitals, and public health units. The PHO HCV Ab and HCV RNA test result data were linked at ICES. ICES is an independent, non-profit research institute whose legal status under Ontario’s health information privacy law allows it to collect and analyze health care and demographic data, without consent, for health system evaluation and improvement. All data were accessed for research purposes on October 19^th^, 2023, at which point data from all participants had already been anonymized with a unique identifier code. authors had access to information that could identify individual participants during or after data collection Data for HCV risk factors were obtained, including HIV status, substance use disorder (SUD, injection drug use not recorded), and birth cohort (to capture generations with an elevated HCV risk).[14] HIV positivity was defined using the ICES HIV Database, which identifies HIV infection using a validated algorithm.[15] SUD was defined by a diagnostic code for drug dependence from the Discharge Abstract Database, the National Ambulatory Care Reporting System database, and the OHIP billing claims database. The diagnostic codes used to identify HIV and SUD are presented in **Table S1**. The Registered Persons Database was used to define eligibility and birth cohort. Estimates of the annual population of Ontario aged 18-80 were found using the ICES POP database, which houses provincial and national population size estimates carried out by Statistics Canada. These datasets were linked using unique encoded identifiers and analyzed at ICES.

The annual count of HCV Ab, Ab+, RNA, and RNA+ tests in the PHO dataset were plotted to assess the impact of these HCV testing trends on each estimation method. To limit uninformative HCV testing data, only the earliest of consecutive HCV RNA+ tests from one individual were kept, and HCV Ab+ tests were removed if an individual had previously tested HCV Ab+.

### HCV incidence estimation methods

Four methods for estimating annual HCV incidence were considered. The methods differ in how they define the at-risk cohort and how they define incident HCV infection. The four methods will be referred to as the population-based method, the test-negative method, the RNA-based method, and the antibody-inclusive method. **Table 1** provides an overview of each estimation method, including the at-risk cohort (denominator) HCV case definition (numerator), and a description of the incidence measurement.

**Table 1.**
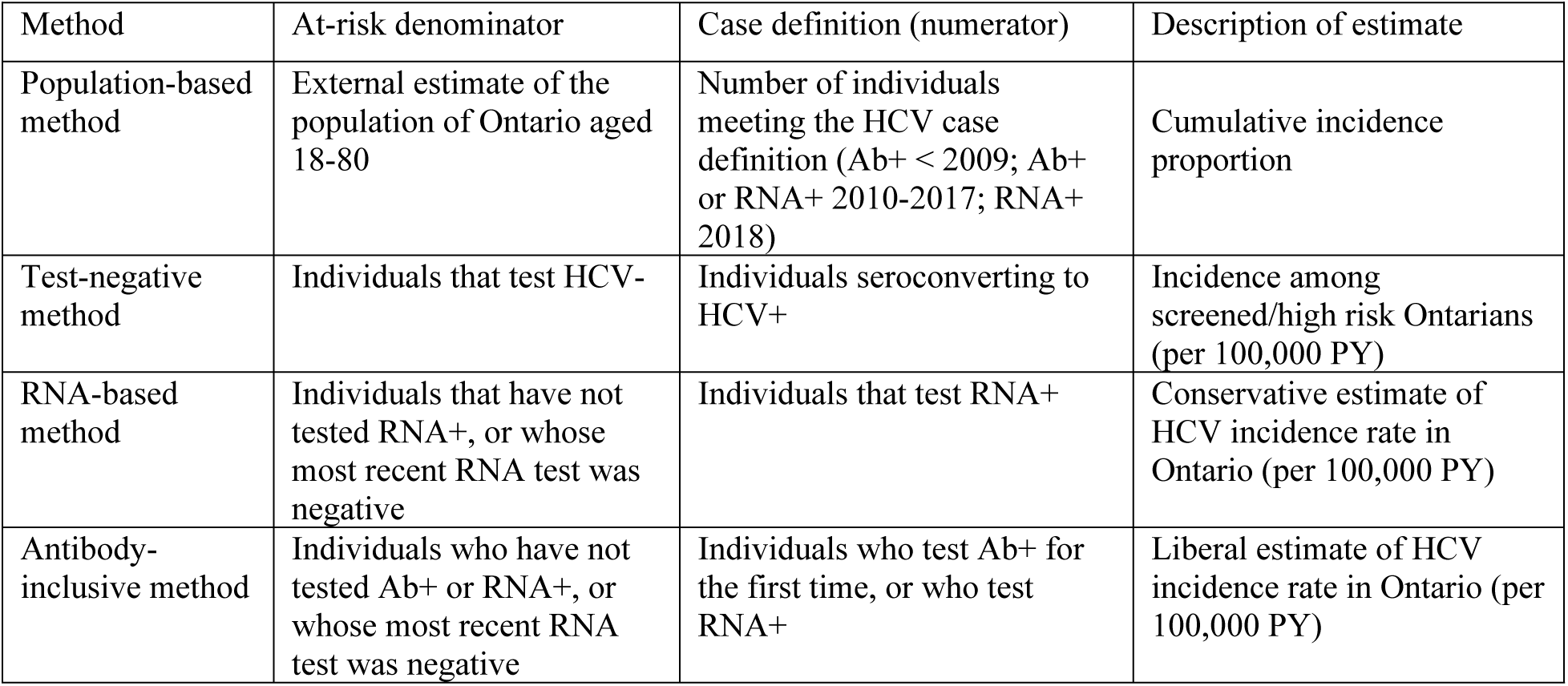
Description of four methods for estimating HCV incidence using extant testing data.

**The population-based method** estimated the annual cumulative incidence of HCV by dividing the number of reported HCV cases in Ontario each year by the population of Ontario aged 18-80 during that year. The HCV case definitions used in this method adhered to the HCV case definitions used by PHO each year; the case definition used by PHO was an HCV Ab+ test result prior to 2009, an HCV Ab+ or RNA+ test result from 2009 to 2017, and an HCV RNA+ test result for 2018.[2, 16] The case count for each year was the number of individuals meeting the HCV case criteria in that year for the first time (e.g. each individual contributes a maximum of 1 case across all years in this method). The denominator of Ontarians aged 18-80 each year from 1999 to 2018 was identified using population estimates from Statistics Canada (the ICES POP database).

**The test-negative method** was restricted to individuals who first tested HCV Ab negative (HCV Ab-) or HCV RNA negative (HCV RNA-). Incident HCV infection was defined as: 1) testing HCV Ab+ or HCV RNA+ for those with a baseline HCV Ab-test result, or 2) testing HCV RNA+ for those with a baseline HCV RNA-test result. HCV+ individuals could re-enter the at-risk population in a later year if they subsequently tested HCV RNA-, indicating infection was cured or cleared. Annual HCV incidence (incidence-density in units of cases per 100,000 person-years) was estimated using Poisson regression.

**The RNA-based method** followed individuals that either: 1) never tested HCV RNA+ or 2) had cleared HCV infection (defined as an RNA+ test result followed by an RNA-test result). Eligibility for the RNA-based method began at time of age 18-80 during 1999-2018, and incident HCV infection was defined as an RNA+ HCV test result.

**The antibody-inclusive method** used the same cohort as the RNA-based method and defined incident HCV infection as the first recorded HCV Ab+ test for an individual, or an RNA+ test.

Regression methods were not required for the population-based method because HCV incidence was estimated by dividing annual case counts by annual population counts. Poisson regression models fit with no covariates were used in the test-negative, RNA-based, and antibody-inclusive methods because these methods used follow-up data, the goal was to estimate HCV incidence, and HCV is a rare event.

### Testing method assumptions and validity

An analysis was carried out to investigate the impact of the population-based method adhering to the changing HCV case definition used by Public Health Ontario (HCV Ab+ <before 2009; HCV Ab+ or RNA+ from 2009-2017, and HCV RNA+ in 2018).[2, 16] The effect of these changes in case definition on the population-based method was determined by estimating annual HCV incidence using HCV Ab+ as the case definition every year, then using HCV RNA+ as the case definition every year.

The generalizability of the test-negative method was evaluated by estimating the association (odds ratios) between established HCV risk factors (HIV, SUD, 1945-1975 birth cohort) and inclusion in the test-negative method cohort, using logistic regression. Additionally, the impact of reassigning the date of HCV infection to be the midpoint between the negative and positive test date was assessed, as this method has been used in previous test-negative cohorts to estimate HCV infection date more accurately.[6, 17]

Potential misclassification in the RNA-based and antibody-inclusive methods was quantified by determining the proportion of first-time Ab+ tests that were unconfirmed by RNA testing, the proportion followed by an RNA+ test, and the proportion followed by an RNA-test.

A final sensitivity analyses was carried out to assess the stability of HCV incidence estimates when time zero was changed from 1999 to 2004. Annual incidence estimates from the sensitivity analysis were divided by those from the original analysis to quantify the effect of changing time zero.

This study received ethical approval from the Queen’s University Health Sciences & Affiliated Teaching Hospitals Research Ethics Board (reference number: 6039130).

## Results

There were 16,118,419 individuals aged 18-80 from 1999-2018. After applying exclusion criteria, the study cohort consisted of 15,757,978 eligible individuals (**Fig 1**). Annual trends of HCV test results from the study cohort are reported in **Fig 2** (annual counts of Ab tests, Ab+ tests, RNA tests, and RNA+ tests). Among the study cohort, there were 1,633,076 HCV Ab tests (83,605 Ab+) and 119,528 RNA tests (69,866 RNA+) used to estimate HCV incidence. While the frequency of Ab+ and RNA+ testing fluctuated moderately over follow-up, Ab and RNA testing increased until a spike in 2015, after which the rate of RNA testing dropped, and the rate of Ab testing continued to rise. **Fig S1** reports these HCV testing trends without removing uninformative HCV tests.

**Fig 1.**
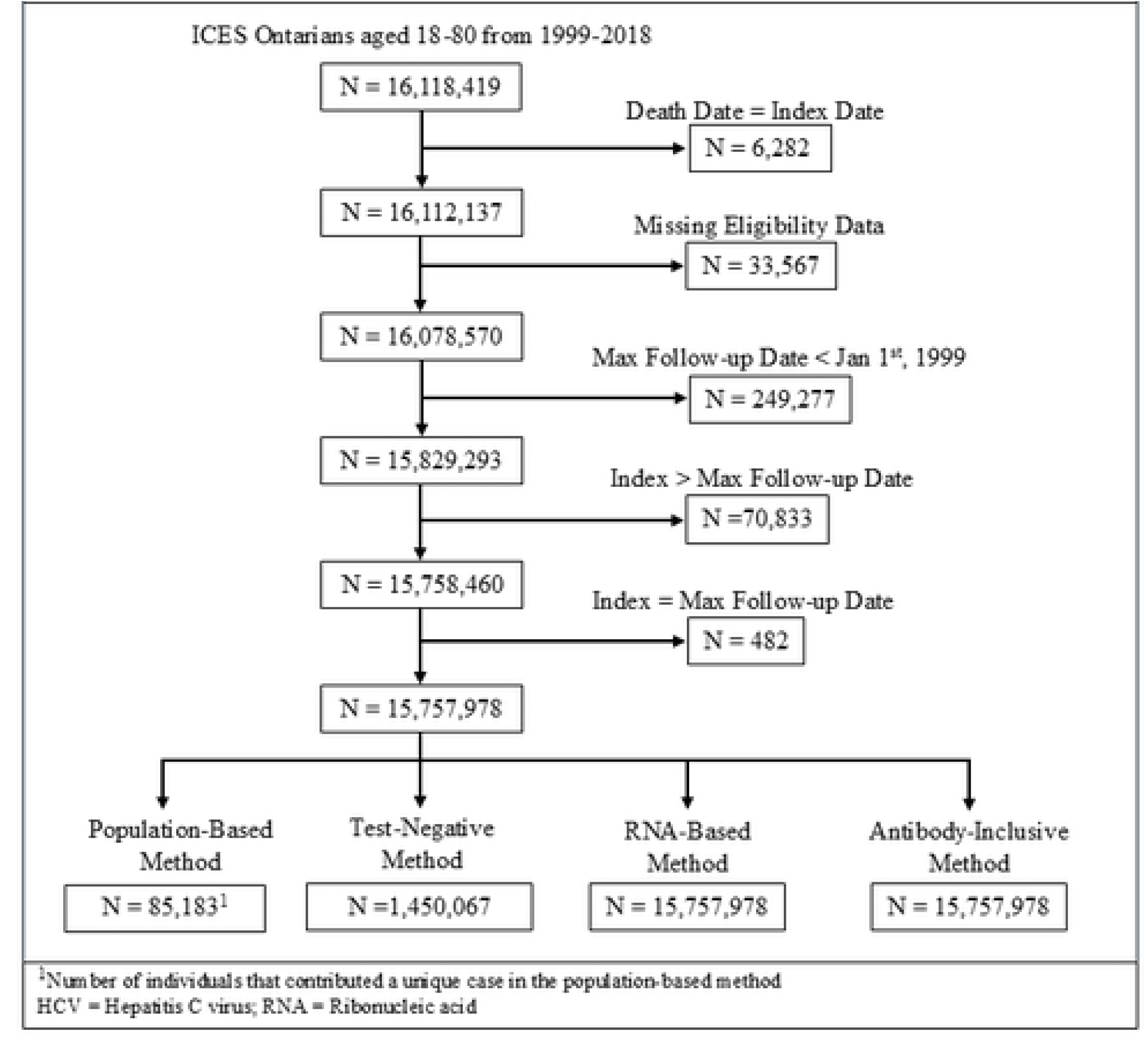
Flowchart of cohort creation for each method used to estimate HCV incidence.

**Fig 2.**
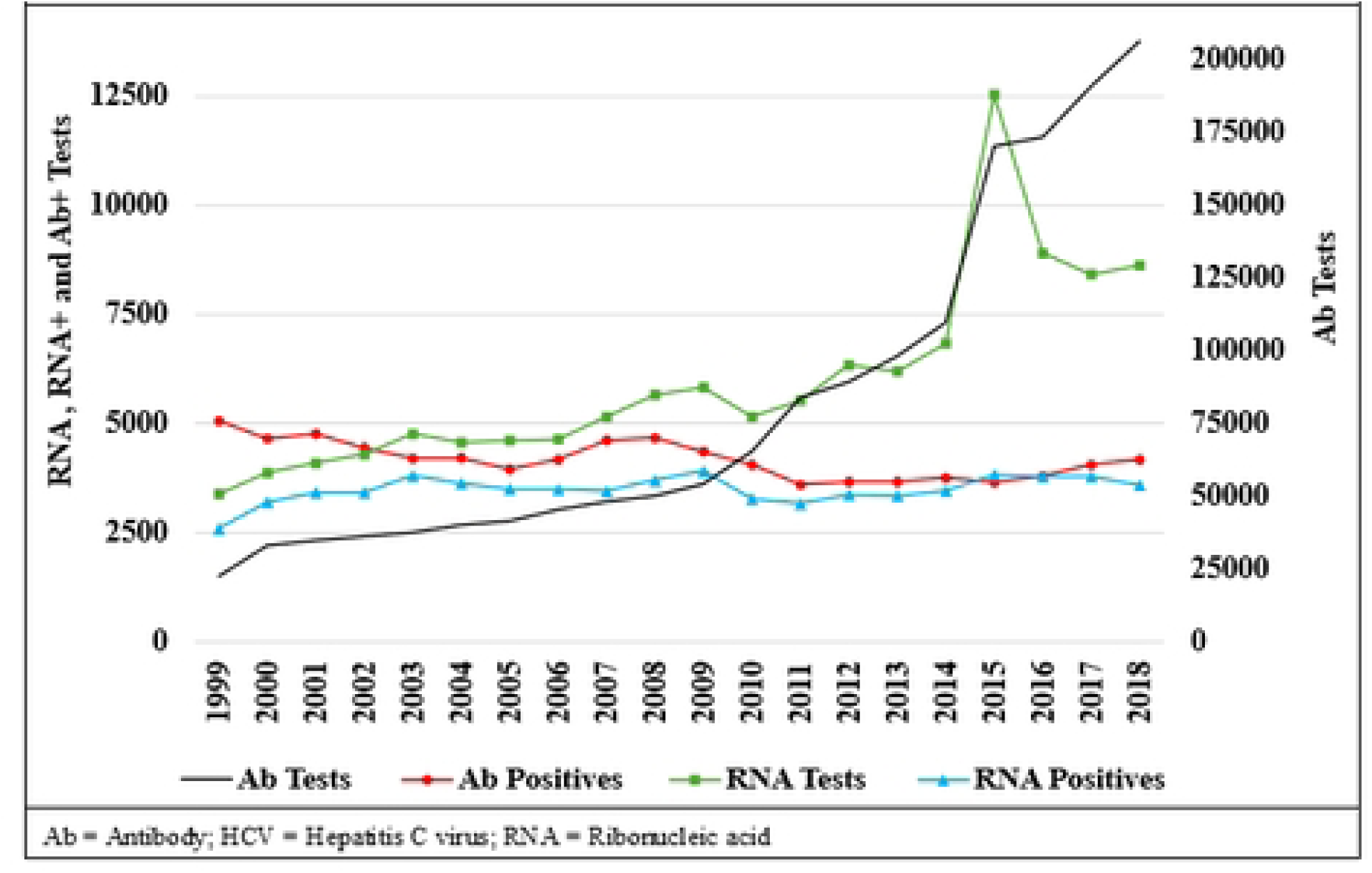
Annual trends of HCV Ab test, Ab+ tests, RNA tests, and RNA+ tests from individuals in the study cohort.

### HCV incidence estimates

HCV incidence estimates from 1999 to 2018 for each method are shown in **Fig 3**. Population-based method estimates of HCV incidence per 100,000 person-years (PY) declined from 59.8 in 1999 to 39.5 in 2011 (with an increase around 2009), remaining stable until a sharp decrease in 2018 to 19.8. Test-negative method estimates were higher than the other methods, and declined from 819.7 cases per 100,000 PY in 1999 (with a spike in 2007), then plateaued at ∼135 cases per 100,000 PY after 2011. RNA-based method estimates fluctuated around 30 cases per 100,000 PY, and antibody-inclusive method estimates declined from 69.5 cases per 100,000 PY in 1999 to 33.4 cases per 100,000 PY in 2011, then slowly rose until reaching 37.5 cases per 100,000 PY in 2018. The mean follow-up time was 5.6 ± 4.9 years for the test-negative method and 14.9 ± 6.3 years for both the RNA-based method and the antibody-inclusive method (follow-up data wasn’t used in the population-based method).

**Fig 3.**
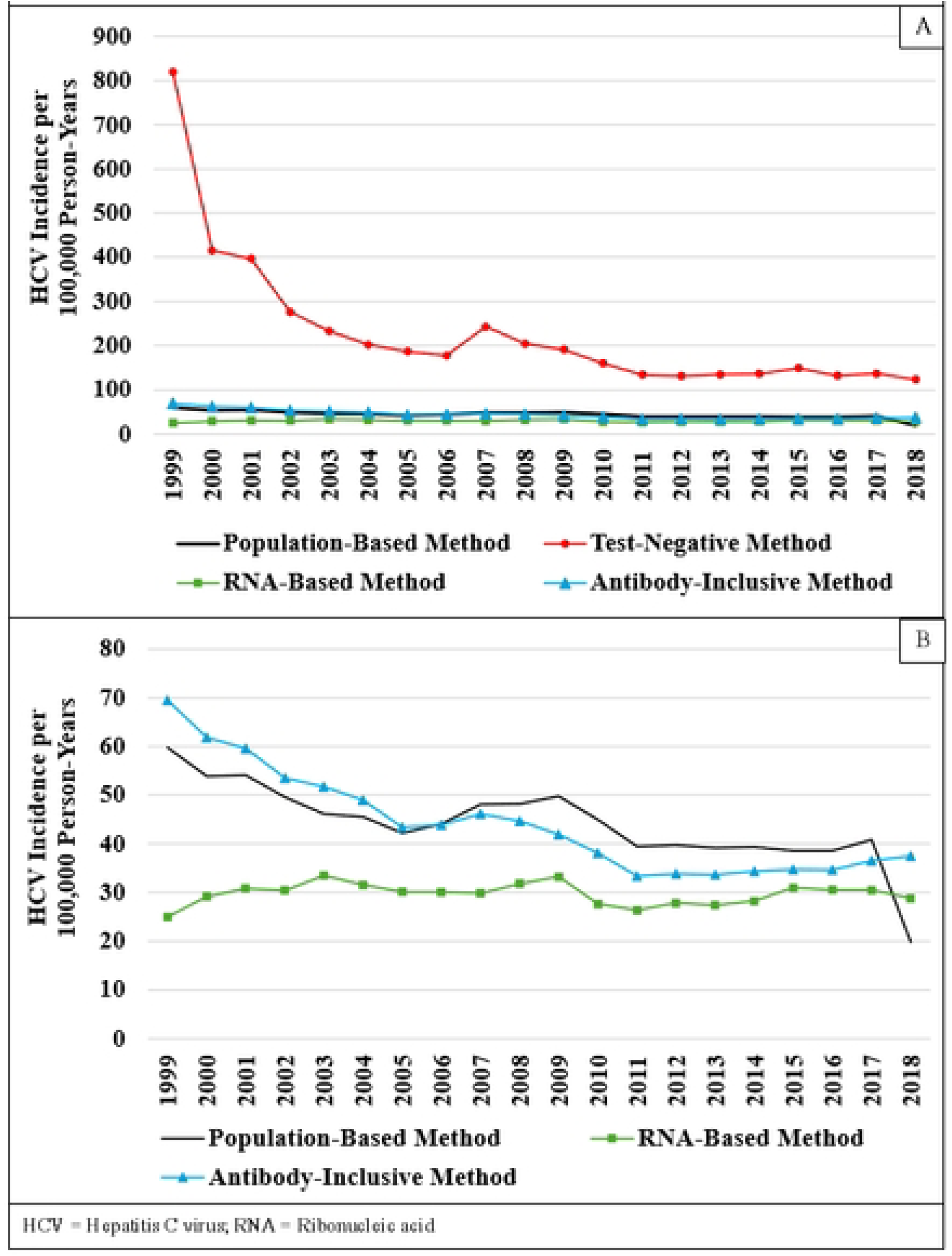
Annual HCV incidence estimates for all estimation methods (Panel A) and the same data without plotting the test-negative method estimates (Panel B)

### Method assumptions and validity

**Fig 4** reports population-based method estimates when using Ab+ testing to define HCV cases every year, and when using RNA+ testing to define HCV cases every year. The incidence estimates from the Ab+ analysis were lower than the original analysis from 2009 onward, except for the 2018. Estimates from the RNA+ analysis were lower than the original in all years (except 2018), and became similar to the Ab+ analysis in 2015 and 2016.

**Fig 4.**
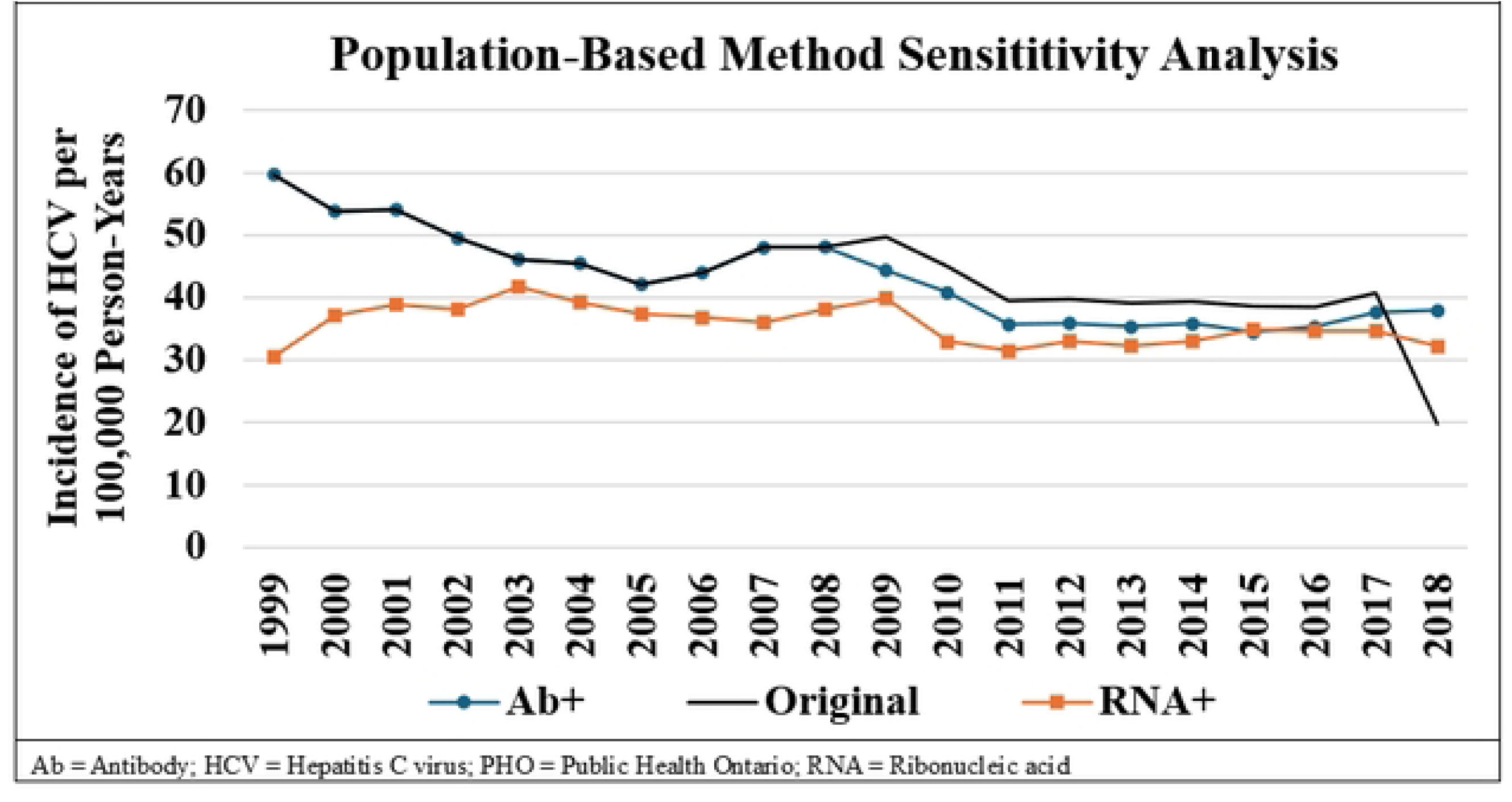
Population-based method HCV incidence estimates, defining HCV cases using the PHO HCV case definition every year (black), using HCV Ab+ every year (blue), and using HCV RNA+ each year (orange)

The odds of being included in the test-negative method cohort were 10.5-fold higher for those positive for HIV (Odds Ratio (OR)=10.5; 95% Confidence Interval (CI): 10.2, 10.7), 2.8-fold higher for those positive for SUD (OR=2.77; 95% CI: 2.76, 2.78), and 4% lower for those born in the 1945-1975 birth cohort (OR=0.964; 95% CI: 0.960, 0.967). Reassigning HCV infection date to be the midpoint between the HCV- and HCV+ test date inflated HCV incidence estimates early in follow-up, and diminished estimates later in follow-up (**Fig 5**).

**Fig 5.**
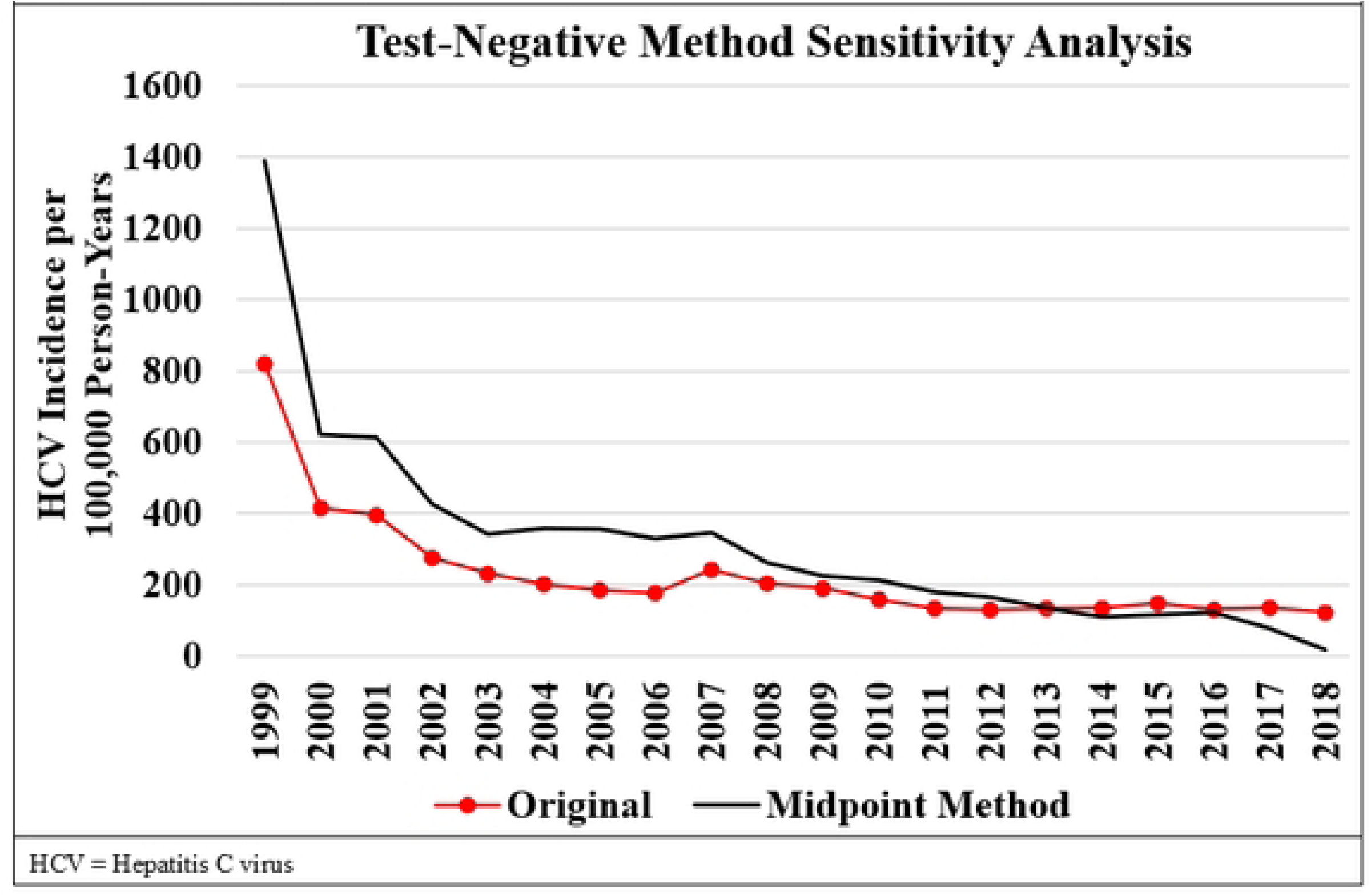
Test-negative method HCV incidence estimates from the original analysis (red), and when HCV infection date was defined as the midpoint between the HCV+ test and the most recent HCV- test (black)

There were 83,605 first-time HCV Ab+ test results in the present study. Of these, 50,921 (61%) were followed by a positive HCV RNA test result, 14,098 (17%) were followed by a negative HCV RNA test result, and 18,586 (22%) were not followed by an RNA test (e.g., unconfirmed HCV Ab+ tests).

The effect of using 2004 as time zero rather than 1999 on HCV incidence estimates are reported in **Fig 6**. Changing time zero had a moderate effect on the population-based method, resulting in higher estimates early on and lower estimates later in follow-up; a much more severe effect of the same pattern was observed in the test-negative method estimates. Changing time zero to 2004 had only a minor impact on the HCV incidence estimates of the RNA-based method, while the antibody-inclusive method consistently had higher estimates after this change.

**Fig 6.**
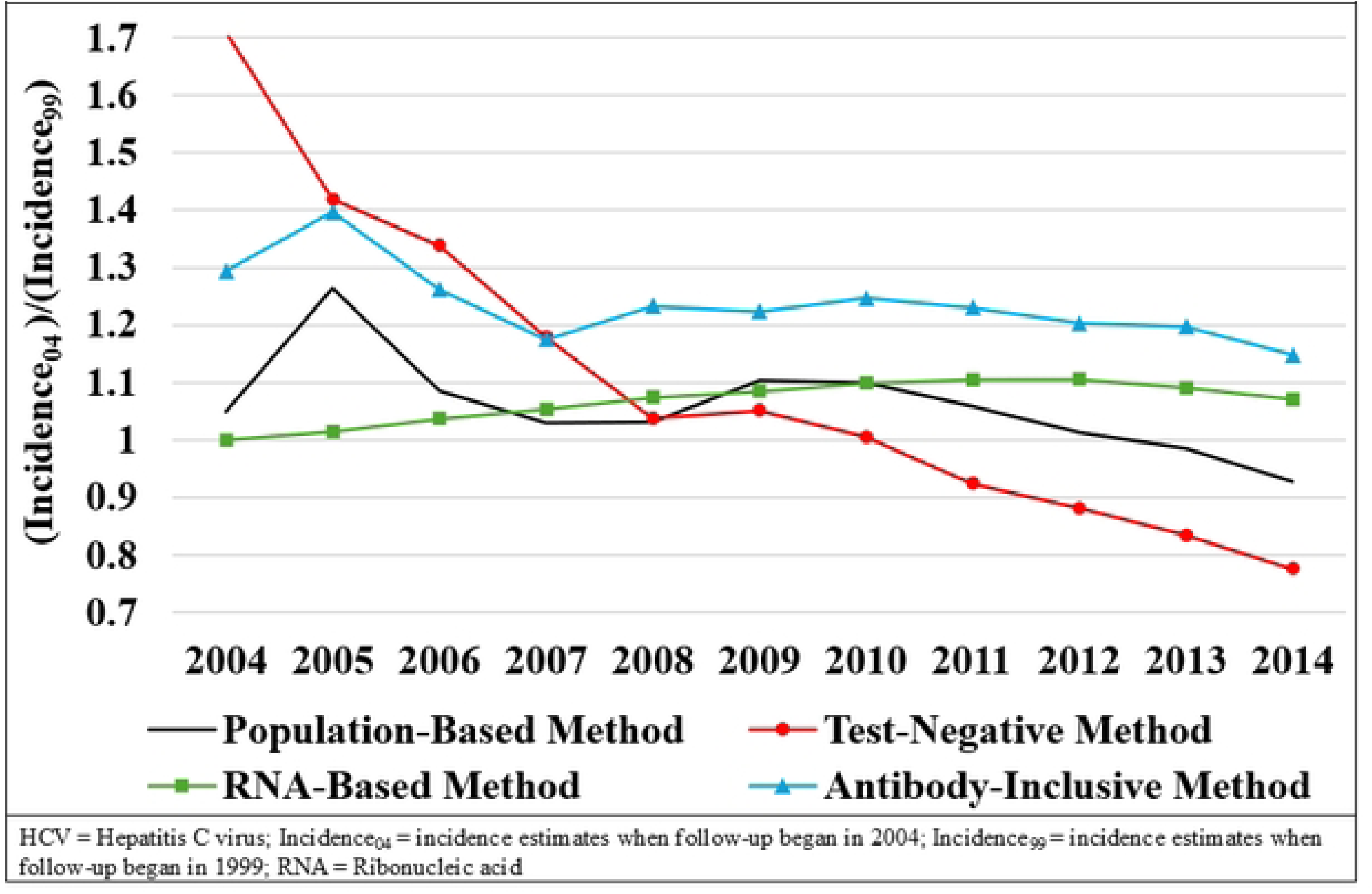
Impact of changing the follow-up start from 1999 to 2004 on each method, reported as the ratio of the HCV incidence estimates with a 2004 start date to estimates with a 1999 start date.

## Discussion

Our study is the first to use a population-based cohort to demonstrate four methods for estimating HCV incidence using extant HCV testing data. Primary concerns when using such data include non-completion of the HCV testing cascade, and differentiating incident and prevalent HCV infections while also preserving generalizability.

The general increase in HCV testing over follow-up can be explained by the rise in the number of Ontarians aged 18-80 over follow-up, and likely also an increased number of individuals at risk for HCV due to the rise in opioid use over this time period.[18] More granular trends can be explained by the release of updated HCV management guidelines in 2005 and 2009, while the large increase in testing in 2015 coincided with the inclusion of the 1945-1975 birth cohort in risk-based screening, and the wide availability of Direct Acting Antivirals as an effective and well-tolerated treatment to cure HCV infection.[9, 19–22]

**The population-based method** used simple calculations and only required aggregate annual HCV testing data. Changes in the Ontario HCV case definition greatly impacted population-based method estimates, limiting its generalizability to other regions that had changes to their HCV case definition in different years, including the USA and the European Union.[23–25] Population-based method incidence estimates dropped in 2018, which can be explained by the fact that the case definition for incident HCV infection was changed to HCV RNA+ in 2018, and a large proportion of individuals that tested RNA+ in 2018 tested Ab+ in a previous year. These individuals therefore contributed an HCV case before 2018 (when Ab+ was part of the case definition), and could not contribute a second case in 2018 when they tested RNA+ (this would consider steps of the HCV testing cascade as 2 separate infection events).

**The test-negative method** was the most internally valid because all HCV cases seroconverted from HCV- to HCV+ while under observation, providing strong evidence that these cases were incident infections. The generalizability of test-negative method incidence estimates was likely limited due to a higher prevalence of HIV and SUD, as these are risk factors for HCV and indications for risk-based HCV screening.[8]

The cohort size for the test-negative method started very small because the cohort each year only consisted of individuals that tested HCV- in that year or a prior year (**Fig S2** plots the annual cohort size and case count for each method). A small cohort size early in follow-up likely explains why HCV incidence estimates for the test-negative method were implausibly high early in follow-up, but began to stabilize over time. This likely also explains why the test-negative method was more susceptible to changing time zero from 1999 to 2004 compared to the other methods.

The test-negative method had the advantage of being able to assign the date of HCV infection to be the midpoint between HCV- and HCV+ testing. While the true date of infection must fall in this window, the clinical relevance of choosing the midpoint relies on the assumption that HCV infections occur halfway between HCV- and HCV+ tests, on average. This change inflated HCV incidence estimates earlier in follow-up and diminished them later in follow-up. This is because cases were only reassigned from later years to earlier years; 1999 gained HCV cases while losing none, and the ratio of lost cases to gained cases increased for each year of follow-up until 2018 when HCV cases were only lost. It is therefore important to recognize that when HCV testing data is only available for a specific window of time, reassigning infection dates to the midpoint between an HCV- and HCV+ test could impact interpretation of trends in HCV incidence.

**The RNA-based method** used a generalizable cohort, was most resistant to changing time 0, and prioritized specificity of HCV classification. In the absence of unconfirmed HCV Ab+ testing, the RNA-based method would likely be the best approach for using extant testing data to obtain generalizable HCV incidence estimates while limiting misclassification. However, HCV incidence was underestimated because 22% of HCV Ab+ tests were unconfirmed by an RNA test, and a proportion of these individuals would have tested RNA+. This proportion may have been large given that most untreated HCV infections become chronic, and HCV treatment cannot be prescribed without testing HCV RNA+.[5, 8, 11] This is further supported by the fact that among HCV Ab+ tests in the present study that *were* followed by an RNA test, it was much more common for this RNA test to be positive (N = 50,921) than negative (N = 14,098). It must also be noted, however, that individuals who do not receive follow-up RNA testing differ in from those that do (e.g., sex, drug use, HIV status).[10]

**The antibody-inclusive method** prioritized sensitivity by including all first-time HCV Ab+ test results as cases, which likely led to overestimation of HCV incidence. When estimating HCV incidence in the presence of unconfirmed Ab+ test results, the antibody inclusive method is a liberal upper bound that assumes all of these unconfirmed Ab+ tests would have tested RNA+ and the RNA-based method is a conservative lower bound that assumes none of them would have tested HCV RNA+. While in theory it would be ideal for the antibody-inclusive method to only include Ab+ tests as cases if they were not followed by an RNA test, this would condition outcome status on a future event and would therefore create immortal-time bias. It was therefore necessary for the antibody-inclusive method to included Ab+ individuals as cases regardless of whether there was data indicating they went on to test RNA-. Nonetheless, HCV incidence estimates from the antibody-inclusive method closely resembled estimates published by PHO between 1999 and 2018 (**Fig S3**), although these PHO estimates use extant HCV data and are therefor also affected by the biases considered in this study.[4, 26–28]

The present study compared four methods for estimating the incidence of HCV using extant HCV testing data. While each method has limitations, they are all useful depending on the priorities of the analysis. The population-based method is suitable in the absence of follow-up data, and the test-negative method is suitable when internal validity of HCV incidence estimates is prioritized over generalizability. When the intent is to generalize HCV incidence estimates to a population beyond those offered risk-based HCV screening, the RNA-based method should be chosen if specificity is a priority, while the antibody-inclusive method should be chosen if sensitivity is a priority.

## Acknowledgements

This study was supported by ICES, which is funded by an annual grant from the Ontario Ministry of Health (MOH) and the Ministry of Long-Term Care (MLTC). This document used data adapted from the Statistics Canada Postal Code^OM^ Conversion File, which is based on data licensed from Canada Post Corporation, and/or data adapted from the Ontario Ministry of Health Postal Code Conversion File, which contains data copied under license from ©Canada Post Corporation and Statistics Canada. Parts of this material are based on data and information compiled and provided by the Ontario Ministry of Health and Statistics Canada. The analyses, conclusions, opinions and statements expressed herein are solely those of the authors and do not reflect those of the funding or data sources; no endorsement is intended or should be inferred.

This study also received funding from: the Natural Sciences and Engineering Research Council of Canada Discovery grant (Y.P.) and the Queen’s Fund for Scholarly work (S.B.).

## Data availability statement

The dataset from this study is held securely in coded form at ICES. While legal data sharing agreements between ICES and data providers (e.g., healthcare organizations and government) prohibit ICES from making the dataset publicly available, access may be granted to those who meet pre-specified criteria for confidential access, available at www.ices.on.ca/DAS. The full dataset creation plan and underlying analytic code may be available from the authors upon request, understanding that the computer programs may rely upon coding templates or macros that are unique to ICES and are therefore either inaccessible or may require modification. Analysis was carried out using SAS Enterprise Guide, Version 8.3 (SAS Institute Inc., Cary, NC, USA).

